# A Review of Point-of-Care Devices for Blood-Testing Towards AI-driven Remote Digital Care, Precision Healthcare and Predictive Medicine

**DOI:** 10.64898/2025.12.13.25340658

**Authors:** Junjia Gu, Hector Zenil

## Abstract

Point-of-care (POC) blood testing enables rapid, decentralized diagnostics with transformative promise, yet its innovation landscape remains poorly mapped. To this end, we focused on features that we believe are key to make progress in areas of precision healthcare and predictive medicine, such as longitudinal data collection and data analytics integration. While no review can be complete, this work attempts to address this gap by analyzing 86 POC blood testing devices worldwide and proposing a unified framework to compare them across technology principles, diagnostic breadth, usability, regulatory pathway, deployment feasibility (via a custom index), and data/AI integration. Electrochemical biosensors were the single largest platform (29.1%), strongly associated with glucose testing (χ^2^=237.8, p<0.001), while spectroscopic and microfluidic systems remained niche due to higher costs and specialized requirements. Regulatory approval skewed toward moderate risk (44.2% FDA II; 27.4% IVDR C), while approval times lengthened with risk class (e.g., IVDR D ≈540 days). A trade-off was observed between usability and panel breadth: tools for home or low-resource settings emphasize simplicity and affordability, whereas clinical systems expand diagnostic range at higher complexity and cost. Deployment feasibility scores favored handhelds, while benchtops were penalized by workflow and capital demands, and microfluidics by consumables. Innovation clusters in North America, Europe, and East Asia reinforce global leadership and disparities.

## 1. INTRODUCTION

Blood testing remains a cornerstone of medical diagnostics, providing essential insights into health status and disease progression. However, traditional laboratory blood tests typically require multiple processing steps, such as sample collection, transport to centralized facilities, and complex analyses, that incur significant time delays, specialized personnel needs, and high costs. These constraints impede timely clinical decision-making, especially in urgent care scenarios or resource-limited settings [1]. In response, point-of-care testing (POCT) and portable devices have gained prominence by effectively “bringing the laboratory to the patient,” enabling rapid, on-site diagnostic results that can accelerate treatment and improve outcomes. The growing demand for accessible diagnostics, driven by expanding healthcare services globally, underscores the significance of POCT, portable and wearable devices – diagnostic tests are essential for identifying patients, guiding therapy, monitoring disease, and evaluating prevention efforts across diverse health interventions [2] but also to enable precision healthcare and predictive medicine.

POCT is an umbrella term that can include portable, handheld and even wearable devices. POCT devices align closely with international health priorities, including the World Health Organization’s ASSURED criteria, which articulate that ideal point-of-care diagnostics should be *Affordable, Sensitive, Specific, User-friendly, Rapid, Equipment-free,* and *Deliverable* to end-users [3]. These benchmarks, originally defined to guide effective testing in low-resource settings, emphasize the importance of accessibility and reliability in decentralized healthcare. Over the past decade, continuous innovation has pushed POCT technologies closer to meeting these goals. Advancements in miniaturized biosensors, microfluidic “lab-on-a-chip” systems, and portable spectroscopic assays have revolutionized the field, yielding ever-smaller devices with performance approaching that of centralized lab tests. At the same time, the proliferation of easy-to-use, network-connected diagnostic devices is integrating POCT into broader eHealth ecosystems, enabling remote patient monitoring and telemedicine applications [4]. This convergence of connectivity and diagnostics reflects a broader trend toward smart, data-driven healthcare delivery. Furthermore, the incorporation of artificial intelligence (AI) and wearable technology is opening new frontiers in continuous health monitoring and personalized medicine. For instance, continuous glucose monitoring systems augmented by AI algorithms can analyze dynamic blood sugar data in real-time, providing individualized feedback and proactive management strategies for diabetes care [5]. Such examples illustrate how next-generation POCT platforms are not only rapid and convenient but also increasingly intelligent and preventive in their approach to healthcare management.

Despite these promising developments, a critical review of the literature reveals a fragmented analytical landscape. Many recent studies and reviews have remained siloed – focusing on specific diseases, device types, or technologies – rather than examining point-of-care devices in a comprehensive, comparative context. For example, some analyses have been confined to nucleic-acid-based testing platforms [6] or to particular assay formats like lateral flow immunoassays [7]. There is a notable lack of systematic evaluations that span the diverse spectrum of blood testing devices available worldwide, across multiple dimensions of innovation. In particular, few works have holistically compared different technological principles (e.g. electrochemical sensors vs. microfluidic devices), varied diagnostic applications (from chronic disease management to acute infection testing), distinct regulatory pathways and classifications, or key usability factors such as affordability, portability, and training requirements. Integration capabilities – including the presence of wearable features or AI-driven analytics – and deployment complexity in real healthcare settings are seldom jointly assessed. Moreover, the global distribution of innovation in this sector (for instance, disparities between high-income and low-income regions in device availability) remains under-characterized in existing research. This gap in the literature points to the need for a unifying analysis that can map the current landscape and identify where opportunities and challenges lie across different fronts of technology and implementation.

The present study is motivated by this clear knowledge gap. In this project, we undertake an extensive multi-dimensional evaluation of 86 representative POCT blood testing devices currently on the market (or in late-stage development), explicitly aiming to provide the first broad analytical mapping of this landscape. Our approach integrates rigorous data analytics, quantitative comparisons, and in-depth literature synthesis to capture a wide array of device attributes and performance metrics. By categorizing devices according to their detection principles, diagnostic targets, usage environments (from clinical laboratories to home settings), and regulatory designations, we establish a framework for cross-comparison that has been missing in prior analyses. We further introduce standardized indicators – such as training difficulty, maintenance needs, and health-system integration capability – to objectively score and compare the deployment feasibility of different device types. Through this comprehensive methodology, the study examines not only technological and clinical features, but also user-centric and system-level considerations (including interoperability, data privacy, and sustainability), aligning our analysis with current regulatory standards and engineering best practices (e.g. FDA and EU IVDR guidelines).

Ultimately, the goal of this research is to fill the literature gap with a holistic overview of the blood testing device innovation landscape. We seek to highlight prevailing trends, pinpoint unmet needs or innovation gaps, and discuss the challenges – technical, regulatory, and ethical – that influence the adoption and scalability of POCT devices. The insights derived from this work are intended to inform a broad audience of stakeholders: clinicians exploring point-of-care solutions, developers and engineers designing the next generation of diagnostics, policymakers and global health organizations strategizing on diagnostic access, and investors or industry leaders making decisions in this evolving market. By elucidating the current state-of-the-art and its deficiencies, our findings aim to support evidence-based decisions and encourage inclusive, sustainable innovation in blood diagnostics. In sum, this study provides a timely and comprehensive “analytics of innovation” in the POCT device landscape, offering a foundation for future research and development efforts to build upon in advancing global healthcare equity and effectiveness.

## 2. METHODS

This study aims to systematically assess the current state of innovation and application potential of Point-of-Care (POC) blood testing devices, which have become an important tool to address the needs of chronic disease management and health monitoring in the context of an aging population due to their rapidity, efficiency, and applicability to near-home scenarios. However, there is still a lack of systematic and comprehensive analyses of the technology classification, diagnostic functions, usage scenarios, deployment complexity, and market resources invested in POC devices globally.

This study identified 86 representative commercially available or late-developed POC blood testing devices, and analyzed in detail the technical principles, wearable features, testing panel coverage, regulatory risk levels, and the time and financial resources required for market launch of each type of device by constructing a multi-dimensional classification framework. In addition, the study developed a device-specific deployment capability scoring system to quantify the feasibility of deploying different device types in real healthcare environments, using training difficulty, maintenance complexity, and healthcare system integration capability as assessment criteria.

Based on the above analysis, this study presents the technology distribution pattern, cost structure and global innovation hotspot maps of POC devices using a multi-dimensional statistical visualization method, forming a complete, clear and easily accessible information map of POC devices, which can provide an intuitive and reliable decision-making basis for clinical decision makers, health technology developers, investment institutions and regulatory authorities.

This study adopts a cross-sectional research design approach to conduct a systematic technology landscape analysis of global Point-of-Care (POC) blood testing devices. The unit of analysis for the study is defined as the device itself, and the sample scope is POC medical devices in the blood testing category that are either on the market or in the late stages of development between 2015 and 2025 to ensure the timeliness, representativeness, and applicability of the study.

In order to clarify the basis for the selection of the study population and to guarantee the reproducibility of the study, strict and clear inclusion and exclusion criteria were set, as shown in Table 2.1:

**Table 2.1.** Inclusion and Exclusion Criteria.

| No. | Inclusion Criteria | Exclusion Criteria |
| --- | --- | --- |
| 1 | Devices that were launched between 2015–2025 or are in late-stage development (e.g., have submitted regulatory applications or completed clinical validation). | Devices are still in prototype design, animal testing, or early-stage development, lacking clear regulatory progress and market information. |
| 2 | Devices with clear and publicly available technical specifications or Instructions for Use (IFU). | Devices lacking publicly available specifications, regulatory information, or detailed product documentation. |
| 3 | Devices with clear registration or listing records in at least one major regulatory authority (e.g., FDA, IVDR, or NMPA). | Devices without identifiable registration or listing records in major regulatory authorities such as FDA, IVDR, or NMPA. |
| 4 | Devices classified as Point-of-Care (POC), explicitly designed for near-home, self-testing, or clinical bedside settings, with features such as portability, ease of use, and rapid response. | Devices clearly intended for centralized laboratory settings or dependent on large-scale instrument platforms, not meeting the definition of POC testing. |

After the screening of the above criteria, 86 POC blood testing devices were finally included as research samples in this study, covering seven major technology platforms, including electrochemical biosensors, spectral analyzers, microfluidic chips, fluorescent immunoassay analyzers and cell counters, and covering the main diagnostic functions, such as clinical chemistry, glucose monitoring, and complete blood cell count (CBC). The study covers multiple dimensions such as technology classification, application scenarios, regulatory levels, deployment complexity, economic costs and global distribution of innovation hotspots of the devices to ensure a comprehensive and in-depth assessment and analysis of POC devices.

### 2.3 Data Source Strategy

To ensure comprehensiveness, reliability, and representativeness, this study implemented a systematic, multi-dimensional data retrieval strategy drawing from regulatory databases, commercial sources, academic literature, and official company websites.

Three main approaches were used. First, academic literature and company websites were systematically searched for detailed information on device specifications, testing parameters, application scenarios, maintenance, training needs, and integration capabilities. Literature searches in PubMed, Google Scholar, and Web of Science used keywords such as *“POC blood testing,” “wearable biosensor,” “microfluidic diagnostics,”* and *“AI-integrated diagnostics”*. Sources included technical reviews, clinical studies, and patents. Company websites were accessed manually to download official Technical Specification Sheets (TSS) and Instructions for Use (IFU). All 86 devices were collated and cited in standardized form [8–93].

Second, regulatory databases were queried for device classification, registration status, and approval timelines. These included the US FDA (510(k), De Novo, GUDID) [94], the EU IVDR [95], and the Chinese NMPA [96]. Searches used device names, manufacturer names, or registration numbers to confirm status one by one.

Third, commercial intelligence sources provided data on market performance, funding, and manufacturer background. Platforms included Crunchbase, PitchBook, and public reports by Grand View Research and MarketsandMarkets. Keywords such as *“POC blood testing device,” “funding,”* and *“market size”* guided searches. A custom Python 3.11 web-scraper of publicly available data (using requests, BeautifulSoup, and pandas) extracted structured fields such as funding amounts and normalized currencies to USD (via forex-python) for research purposes. The scraper used request throttling and anti-bot protections, with a 10% manual validation sample ensuring ≥95% accuracy. Outputs were merged into the Excel dataset for analysis. The complete workflow is summarized in Table 2.2.

**Table 2.2.** Overview of Semi-Automated Funding Data Extraction Process.

| Step | Tool / Library | Key Controls | Output Fields |
| --- | --- | --- | --- |
| **1. URL List Compilation** | Manual search on Crunchbase / PitchBook | Company name + product name<br>cross-verification | url_list.csv |
| **2. HTTP Requests** | requests | User-Agent spoofing;<br>3-second interval<br>between requests | HTML |
| **3. Data Parsing** | BeautifulSoup | Locate “Funding Rounds” section | Funding amount<br>(original currency) |
| **4. Currency Conversion** | forex-python | Record exchange rate<br>timestamp | Funding amount<br>(USD) |
| **5. Data Entry** | pandas | Append date of<br>retrieval | Excel “Funding”<br>column |

Through the above multi-channel and multi-method data acquisition and cross-validation strategies, this study constructed a systematic, traceable and reliable quality dataset to ensure the credibility and academic rigor of the analysis results.

### 2.4 Data Cleaning & Standardization

To ensure accuracy, consistency, and reliability for subsequent statistical analysis and visualization, the dataset underwent systematic cleaning and standardization in Microsoft Excel 365, following defined operational standards to maintain traceability and reproducibility.

Duplicate handling: A unique identifier (device + manufacturer) was generated to detect duplicates. Records were manually reviewed, and the most complete entry was retained.

Unit standardization: Measurement units were converted for consistency. Volumes (cm³, mL, m³) were standardized to cubic decimeters (dm³); mass values (kg, mg) to grams (g). Conditional Excel formulas and manual verification ensured conversion accuracy. Extreme values were flagged and individually validated.

Financial normalization: Device prices and funding amounts (EUR, CNY, etc.) were converted into USD using foreign exchange rates fixed on June 30, 2025. Entries lacking financial data were labeled “Not publicly disclosed” and treated separately in analyses.

Missing values: Supplementary searches were conducted to fill critical gaps. For non-critical numerical fields, median imputation was applied within technology categories when manual searches failed. Critical fields without reliable public data were retained as “Not publicly disclosed” and excluded from relevant statistical tests to minimize bias.

Outlier detection: Boxplot (1.5×IQR) thresholds identified potential outliers in price, volume, and mass. Each case was manually cross-checked. Valid extremes were retained and flagged in results; erroneous entries were corrected or removed.

Documentation: A comprehensive data dictionary captured variable definitions, units, ranges, and all transformations. A summary of cleaning and standardization procedures is provided in Table 2.3.

**Table 2.3.** Overview of Data Cleaning and Standardization Steps.

| Step | Description of Operations | Key Quality Controls |
| --- | --- | --- |
| **1. Duplicate Removal** | Identification and manual verification based on device and manufacturer names | Manual review of all potential duplicates |
| **2. Unit Standardization** | Volume converted to $\text{dm}^3$, mass converted to g using standardized Excel calculations | Manual spot-checking of conversions for accuracy |
| **3. Currency Conversion** | All prices and funding amounts standardized to USD using exchange rates from June 30, 2025 | Exchange rates logged; consistent date reference |
| **4. Missing Value Handling** | Supplementary manual search, median-based imputation, labeling of undisclosed values clearly | Median imputation limited to non-critical numerical fields |
| **5. Outlier Detection** | Boxplot method ($1.5 \times \text{IQR}$) applied to identify potential outliers in price, mass, and volume | Individual manual verification of flagged data |
| **6. Data Documentation** | Structured data dictionary created with clearly defined fields and data transformation procedures | Data dictionary peer-reviewed for completeness |

Through the systematic application of these comprehensive cleaning and standardization steps, the final dataset became robust and suitable for reliable and reproducible downstream classification, scoring, and visualization analyses.

### 2.5 Feature Engineering & Classification

To achieve a structured and comprehensive analysis of POC blood testing devices, this study conducted a multi-dimensional feature engineering process and systematic classification of all devices included in the analysis based on data cleaning and standardization. These feature dimensions cover the Device Use Context, Technology Classification, Test Type, Device Type, AI Capability, and Long-term Data Storage Capability.

Classification criteria and specific classification are based on the following sources: publicly available device registration dossiers from regulatory agencies (e.g., FDA, CE-IVD, NMPA); Instructions for Use (IFUs) and technical white papers issued by device manufacturers; and recent review literature related to POC diagnostic devices. In cases where there was controversy over the specific categorization of an individual device, the research team worked through discussions within the group to reach a consensus to determine its most accurate classification.

For ease of understanding and replication in subsequent studies, the classification criteria and rationale for each dimension are detailed in Table 2.4.

**Table 2.4.**
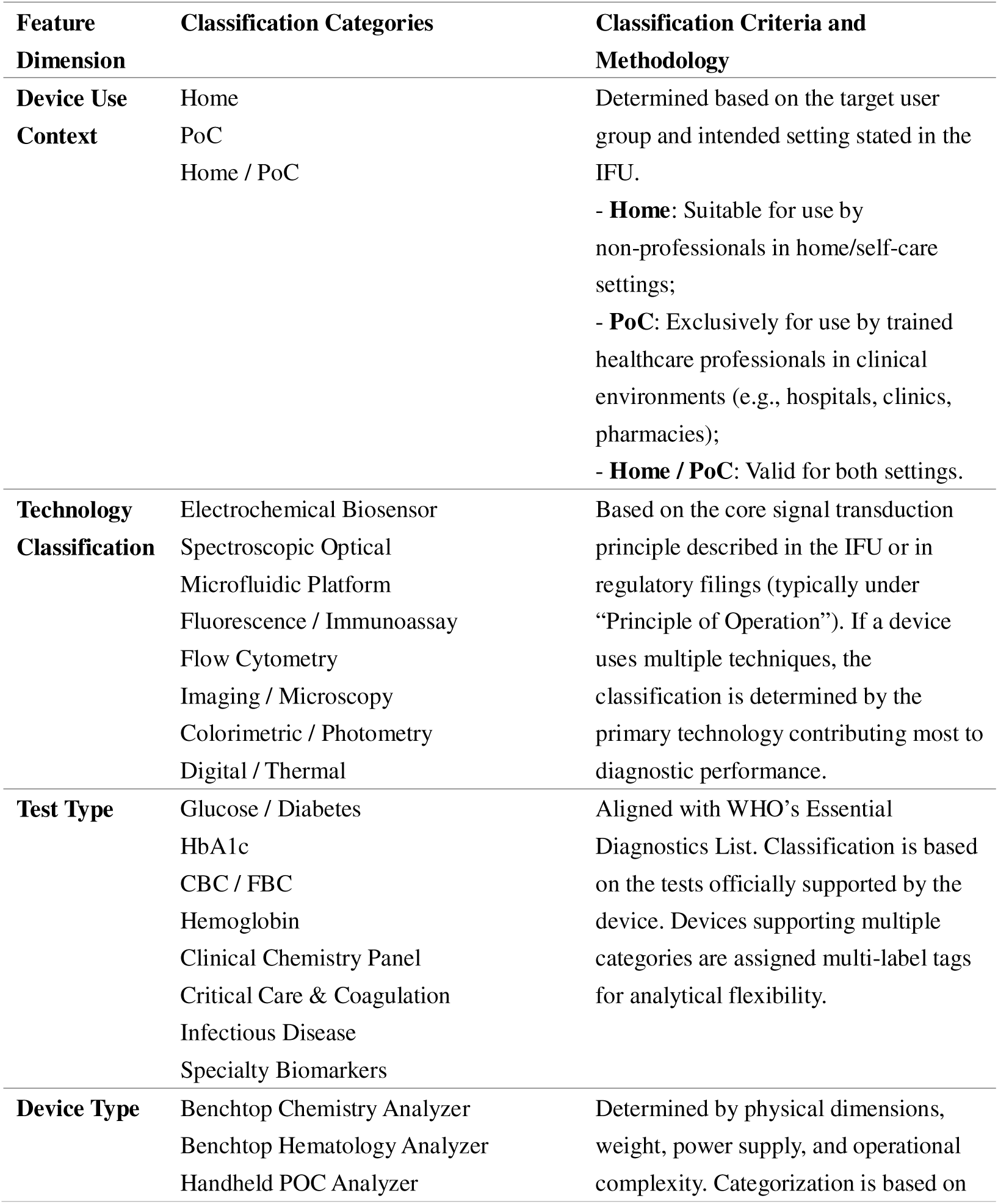
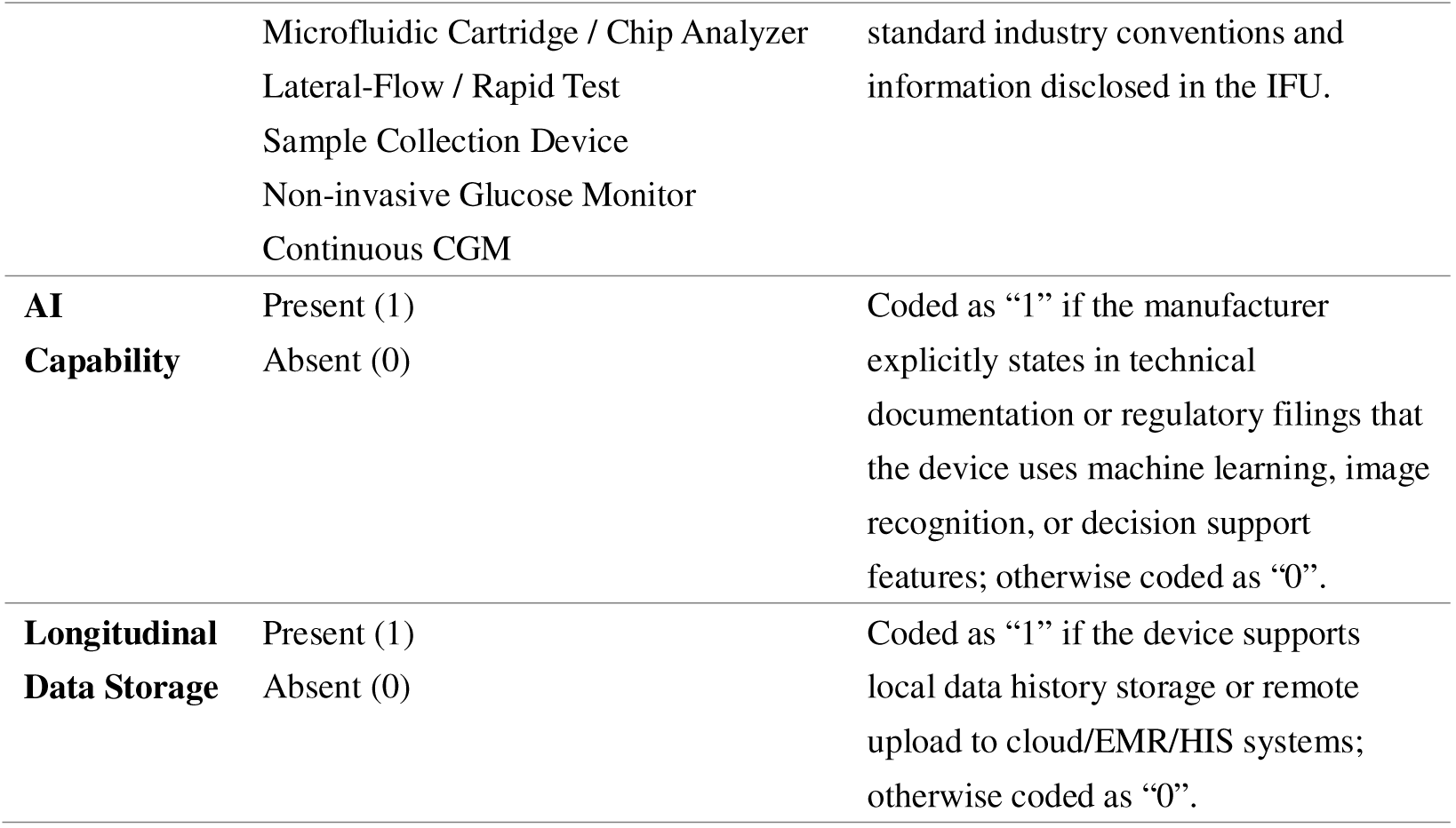
Summary of Feature Dimensions and Classification Criteria.

| **Feature Dimension** | **Classification Categories** | **Classification Criteria and Methodology** |
| --- | --- | --- |
| **Device Use Context** | Home<br>PoC<br>Home / PoC | Determined based on the target user group and intended setting stated in the IFU.<br>- **Home:** Suitable for use by non-professionals in home/self-care settings;<br>- **PoC:** Exclusively for use by trained healthcare professionals in clinical environments (e.g., hospitals, clinics, pharmacies);<br>- **Home / PoC:** Valid for both settings. |
| **Technology Classification** | Electrochemical Biosensor<br>Spectroscopic Optical<br>Microfluidic Platform<br>Fluorescence / Immunoassay<br>Flow Cytometry<br>Imaging / Microscopy<br>Colorimetric / Photometry<br>Digital / Thermal | Based on the core signal transduction principle described in the IFU or in regulatory filings (typically under “Principle of Operation”). If a device uses multiple techniques, the classification is determined by the primary technology contributing most to diagnostic performance. |
| **Test Type** | Glucose / Diabetes<br>HbA1c<br>CBC / FBC<br>Hemoglobin<br>Clinical Chemistry Panel<br>Critical Care & Coagulation<br>Infectious Disease<br>Specialty Biomarkers | Aligned with WHO’s Essential Diagnostics List. Classification is based on the tests officially supported by the device. Devices supporting multiple categories are assigned multi-label tags for analytical flexibility. |
| **Device Type** | Benchtop Chemistry Analyzer<br>Benchtop Hematology Analyzer<br>Handheld POC Analyzer | Determined by physical dimensions, weight, power supply, and operational complexity. Categorization is based on |
|  | Microfluidic Cartridge / Chip Analyzer<br>Lateral-Flow / Rapid Test<br>Sample Collection Device<br>Non-invasive Glucose Monitor<br>Continuous CGM | standard industry conventions and information disclosed in the IFU. |
| **AI Capability** | Present (1)<br>Absent (0) | Coded as “1” if the manufacturer explicitly states in technical documentation or regulatory filings that the device uses machine learning, image recognition, or decision support features; otherwise coded as “0”. |
| **Longitudinal Data Storage** | Present (1)<br>Absent (0) | Coded as “1” if the device supports local data history storage or remote upload to cloud/EMR/HIS systems; otherwise coded as “0”. |

**Table 2.5.**
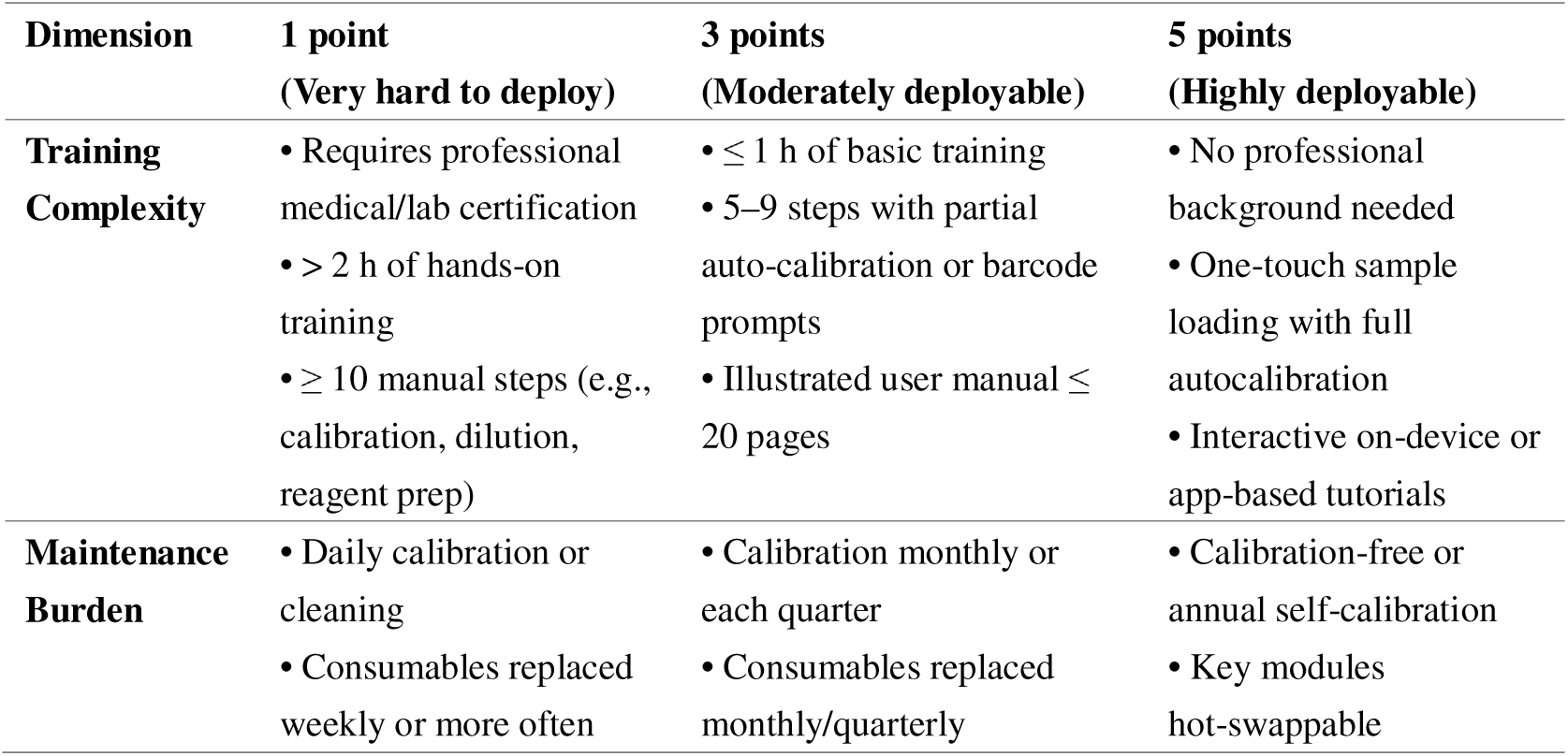
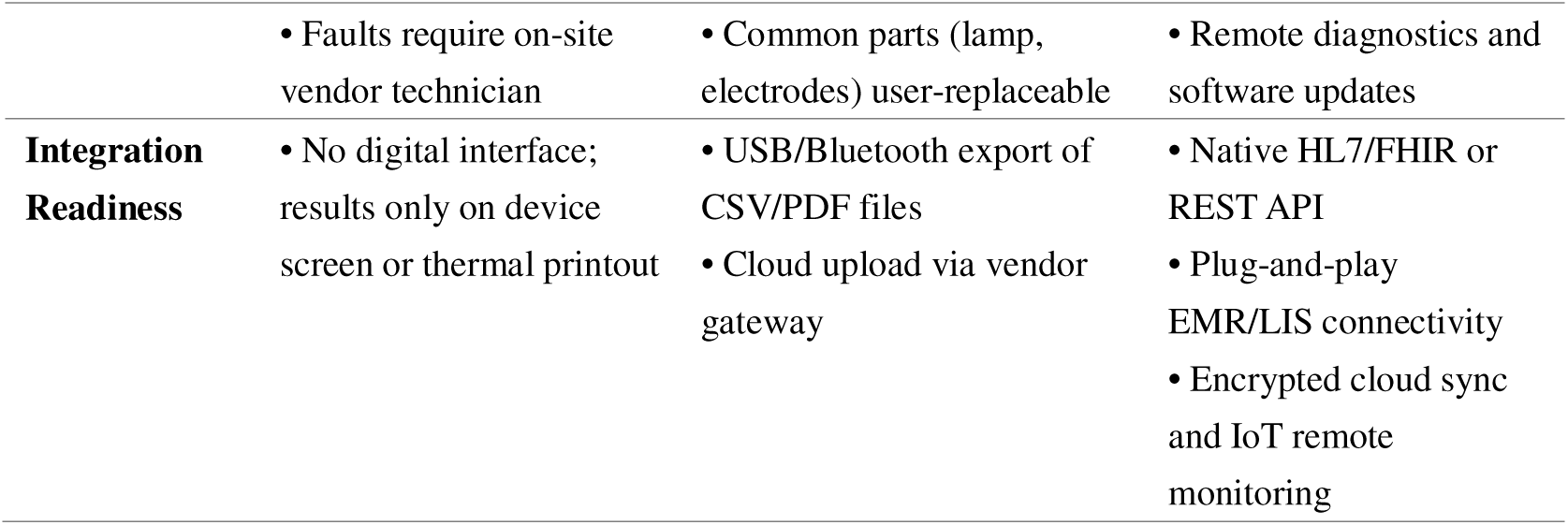
Scoring criteria for the Deployment-Capability Index (1–5 scale)

| **Dimension** | **1 point**<br>**(Very hard to deploy)** | **3 points**<br>**(Moderately deployable)** | **5 points**<br>**(Highly deployable)** |
| --- | --- | --- | --- |
| **Training Complexity** | • Requires professional medical/lab certification<br>• > 2 h of hands-on training<br>• $\geq 10$ manual steps (e.g., calibration, dilution, reagent prep) | • $\leq 1$ h of basic training<br>• 5–9 steps with partial auto-calibration or barcode prompts<br>• Illustrated user manual $\leq 20$ pages | • No professional background needed<br>• One-touch sample loading with full autocalibration<br>• Interactive on-device or app-based tutorials |
| **Maintenance Burden** | • Daily calibration or cleaning<br>• Consumables replaced weekly or more often | • Calibration monthly or each quarter<br>• Consumables replaced monthly/quarterly | • Calibration-free or annual self-calibration<br>• Key modules hot-swappable |
|  | • Faults require on-site vendor technician | • Common parts (lamp, electrodes) user-replaceable | • Remote diagnostics and software updates |
| **Integration Readiness** | • No digital interface; results only on device screen or thermal printout | • USB/Bluetooth export of CSV/PDF files<br>• Cloud upload via vendor gateway | • Native HL7/FHIR or REST API<br>• Plug-and-play EMR/LIS connectivity<br>• Encrypted cloud sync and IoT remote monitoring |

**Table 2.6.** Overview of statistical tools and visual outputs.

| Research objective | Primary visualization | Statistical test |
| --- | --- | --- |
| **Technology–panel distribution** | **Clustered column chart** (platform counts)<br>**Heat-map** (platform × panel frequency matrix) | $\chi^2$: platform × panel |
| **Capital demand & approval time** | **Grouped column chart** (median funding by risk tier & stage)<br>**Boxplot** (approval days by pathway)<br>**Pie chart** (cleared vs uncleared share) | Kruskal-Wallis: funding across three risk tiers |
| **Near-home usability** | **Log-scale boxplot** (volume, weight: Home vs PoC)<br>**Grouped column chart** (price, annual cost: Home vs PoC)<br>**Pie chart** (Home/PoC/Dual share) | Mann-Whitney U: volume, weight |
| **Wearable smart-feature penetration** | **Stacked column chart** (AI + storage combinations by form factor)<br>**Pie charts** (AI coverage; storage coverage) | — (descriptive) |
|  | within wearables) |  |
| **PoC deployment & cost** | **Grouped column chart** (mean DCI sub-scores for four PoC forms)<br>**Grouped column chart** (median annual consumables cost) | Kruskal-Wallis:<br>consumables cost<br>across four forms |
| **Global innovation spread** | **Filled map** (manufacturer count by country, 5-class red gradient) | — (descriptive) |

The above features: engineering and classification steps laid a solid foundation for the subsequent data analysis, and constructed a structured device database that clearly demonstrated the technical and clinical characteristics of each device. With strict and traceable classification criteria, this study not only improves the rigor and reproducibility of data processing and analysis but also provides a reliable reference framework for industry decision makers to classify technologies.

### 2.6 Deployment-Capability Index

To quantitatively assess how readily each POC blood-testing device can be deployed in real-world settings, a three-dimension Deployment-Capability Index was developed, drawing on FDA Human Factors guidance [97] and ISO 62366 usability engineering principles [98]. All scores were assigned by the author based on publicly available technical documents and IFUs; repeated self-scoring of benchmark devices was used to check scale consistency. Where documentation was insufficient, a conservative score of 2 was applied and flagged as “information limited” in the dataset.

**Scoring note.** Each dimension employs integer scores; devices falling between anchor levels may be assigned 2 or 4 points to reflect intermediate performance. Summing the three-dimension scores yields a total DCI of 3–15, with higher totals indicating lower overall deployment effort and superior integration capability.

This DCI framework supplies a quantitative basis for the cost-versus-usability comparisons presented later in the study and offers healthcare organizations a structured reference when planning device selection, staff training, and IT integration strategies.

### 2.7 Statistical Analysis & Visualization

All data collation, statistical tests and graphing were done in Microsoft Excel 365. Frequencies and percentages were calculated using PivotTable for categorical variables; continuous variables, due to significant skewness in the distribution, were uniformly non-parametric (Mann-Whitney U or Kruskal-Wallis) and effect sizes were reported where required (Cramér’s V or η^2^). The spatial distribution is plotted relying on 3D Map. The color palette was uniformly pastel, except for the heat map and the world map, which used a single red gradient from light to dark, to maintain overall visual consistency. The significance threshold was set at α = 0.05.

In the data visualization process, frequency counts were firstly performed for the technology platform and the detection panel, and clustered bar charts were plotted separately; subsequently, the two variables were intersected to generate the two-dimensional frequency matrices, the χ^2^ test was performed and converted to Cramér’s V. The results were visualized as a red-system heatmap to show the enrichment or absence of the technology-detection combinations.

To assess the impact of regulatory risk tiers on the size of financing, the median amount of financing at the early, venture and IPO stages was counted in three tiers: low, medium and high, and the Kruskal-Wallis test was used to compare the differences and report the η^2^ quantitative explanatory rate; the output is presented in a grouped bar chart. The lengths of the main approval paths are characterized by box plots, which are not tested for significance as they are used for descriptive purposes, and pie charts giving the relative proportions of approved versus unapproved devices.

In the near-home availability analyses, volume and mass were log10 transformed and box plotted, and Mann-Whitney U-tests were used to compare Home versus PoC scenario differences; price versus annual consumables is given as median values in grouped bar charts directly as a descriptive control of economic burden, and scenario shares are presented in pie charts.

In terms of intelligence penetration, the frequency of the combination of AI capability and long-term data storage is first aggregated by device form, and a stacked bar chart is plotted, followed by two pie charts showing the AI and data storage coverage of each wearable subcategory; this section is only a descriptive display.

Deployment capability and cost are assessed using the established DCI scores, with the mean values of the three sub-dimensions of training, maintenance and system integration calculated for each of the four PoC forms and plotted in grouped bar charts; the differences in annual consumable costs among the four forms are assessed by the Kruskal-Wallis test and reported as η^2^, also shown in grouped bar charts.

Finally, the manufacturer HQ country counts were imported into a 3D Map to create a 5-isometric segmented red-system coloring map to visualize global innovation hotspots, which is descriptive visualization and does not implement inferential testing. Through the above process, this study provides multi-dimensional visualization results ranging from overall distribution to specific performance differences to spatial patterns in a uniform color scheme and graphical specification while maintaining statistical rigor, providing clear and reproducible quantitative evidence for subsequent discussion and decision-making.

## 3. RESULTS

### 3.1 Categorization by Technological Capabilities and Blood Panels

In this section, the distribution of technology platforms and diagnostic panels used in 86 existing or upcoming POC blood testing devices is systematically presented by classifying and analyzing these devices. The results include the distribution of technology classifications (Fig. 1), the distribution of diagnostic panel types (Fig. 2) and the correlation analysis between technology platforms and testing panels (Fig. 3), to explore the intrinsic relationship between technology choices and diagnostic applications.

**Figure 1:**
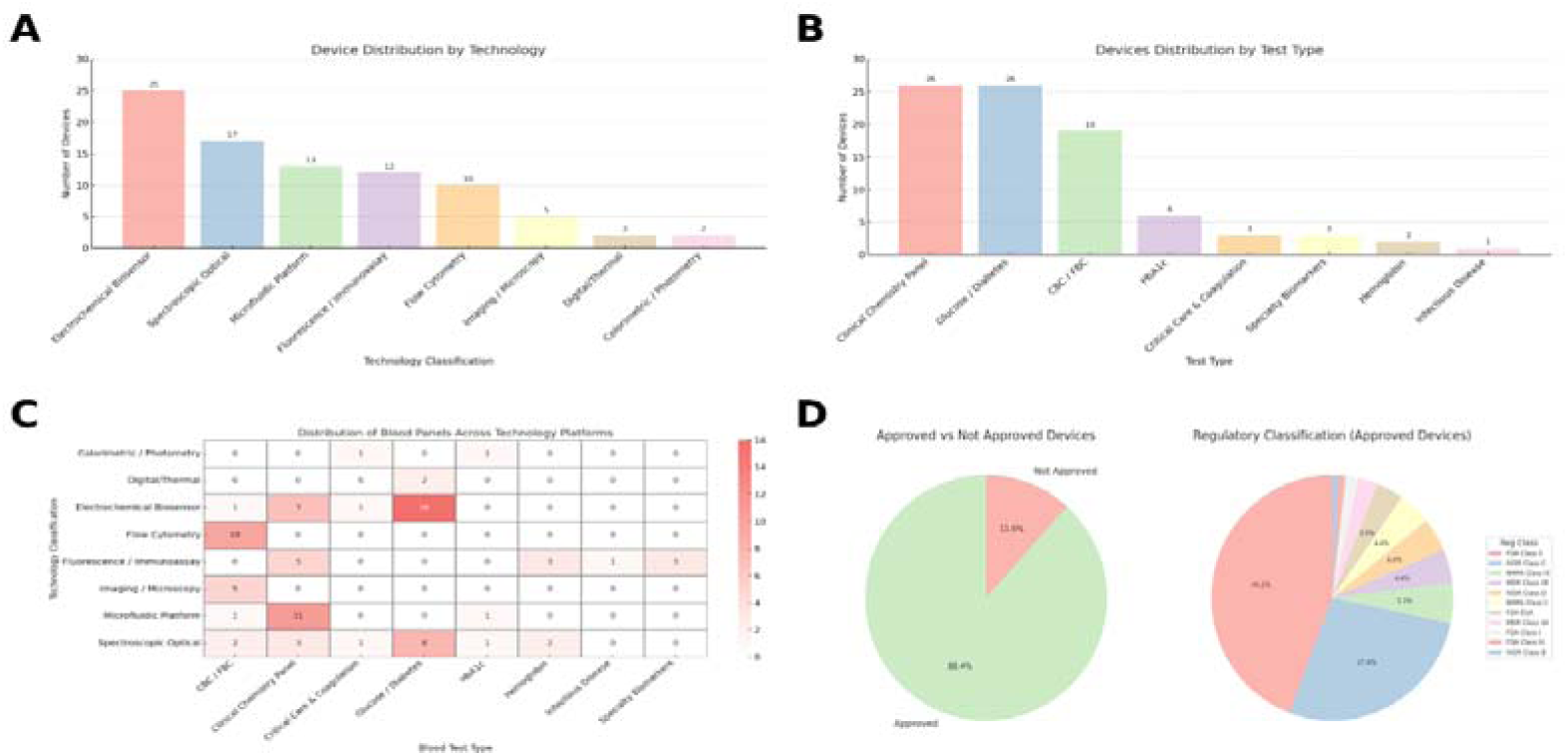
A) Devices Distribution by Technology. B) Distribution of Devices by Blood Test Panel. C) Distribution of Blood Panels Across Technology Platforms. D) Regulatory Approval and Classification Overview

**Figure 2.**
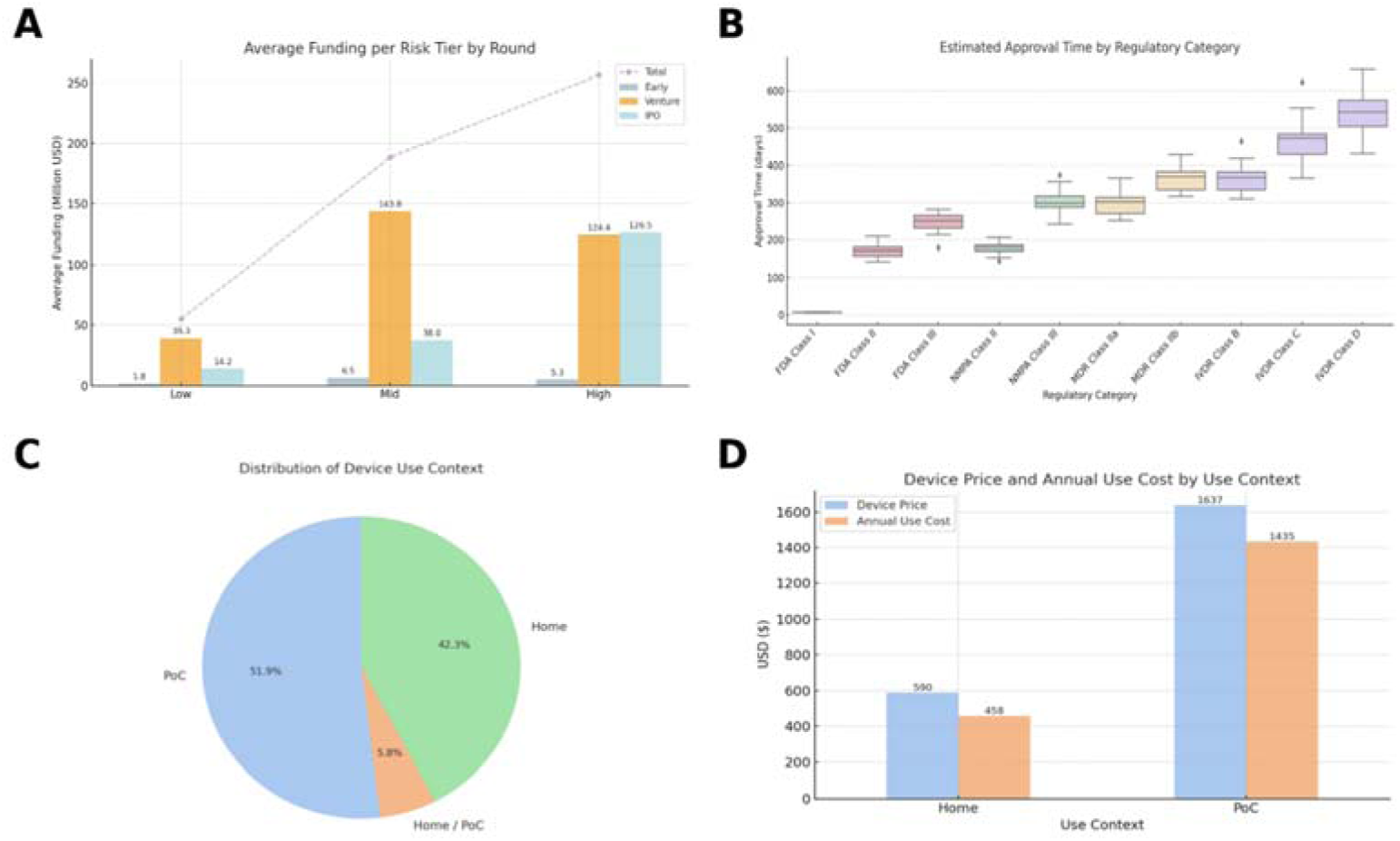
A) Average Funding by Regulatory Risk Tier and Investment Stage. B) Average Time of Regulatory Approval After Submission. C) Device Use Context Distribution, and D) Associated Cost Comparison

**Figure 3.**
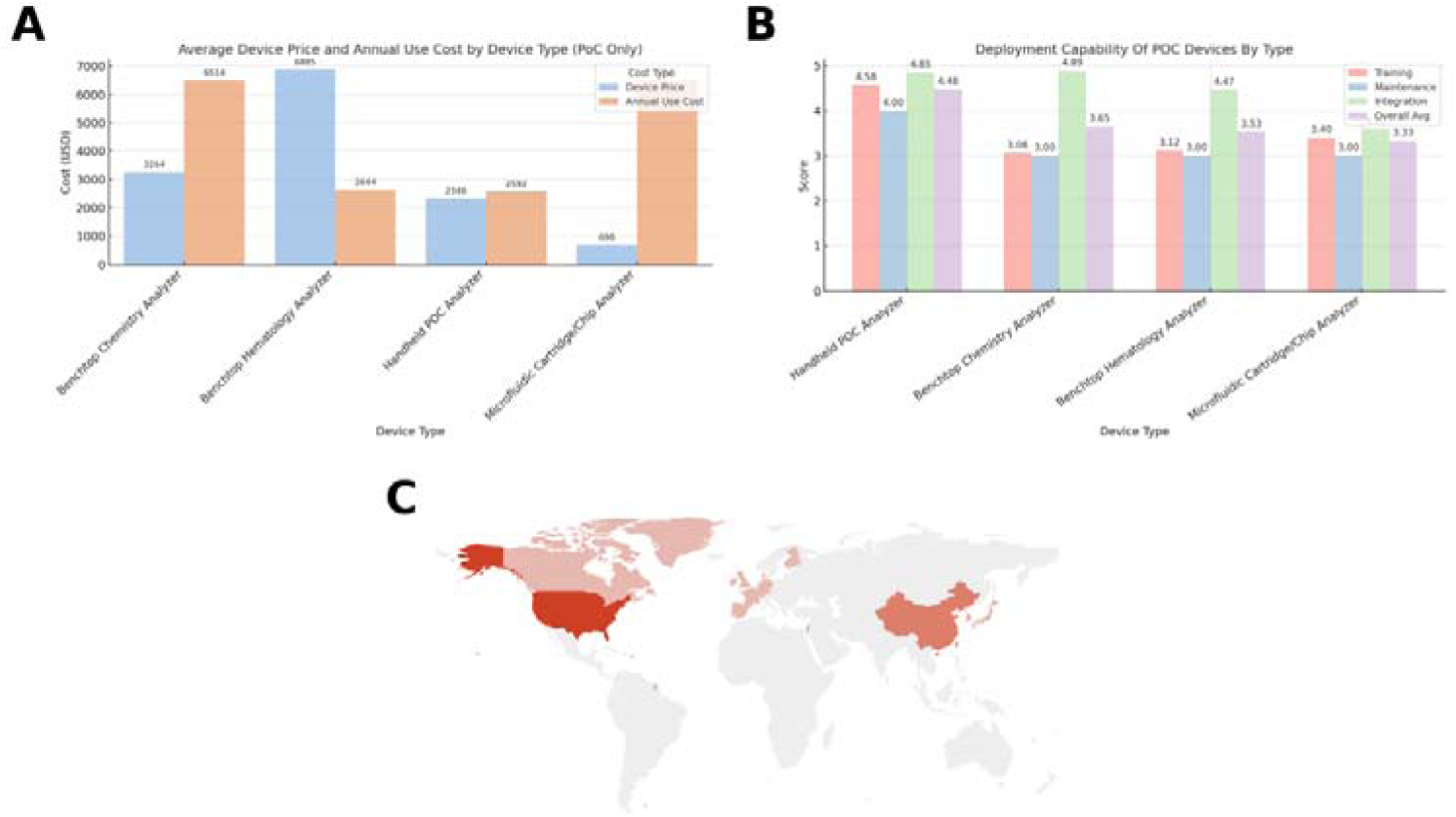
A) Average Device Price and Annual Use Cost by PoC Device Type. B) Deployment Capability Scores of PoC Devices by Type. C) Global Geographic Distribution of Blood Testing Device Developers

Figure 1A shows the distribution of the 86 devices analyzed in terms of technology platforms. The results show that electrochemical biosensor technology is currently the most common technology platform among POC blood testing devices (29.1%, 25 devices), mainly due to its advantages of simplicity of operation, portability and suitability for frequent testing, and it is particularly dominant in the field of blood glucose monitoring. Also featured prominently are spectroscopic optics (19.8%, 17 devices) and microfluidics (15.1%, 13 devices), both of which are gaining traction due to their superiority in multiplexed biochemical assays and miniaturized device development. Fluorescence/immunoassay technology (14.0 per cent, 12 devices) and flow cytometry (11.6 per cent, 10 devices), on the other hand, have a wider range of applications in specialized medical scenarios. Relatively fewer of the more complex technologies such as imaging/microscopy, digital/thermal sensing, and colorimetric/photometric were used, with only 5, 2, and 2 devices, respectively, which may be related to the fact that the devices for these technologies are larger in size, more costly, or less likely to be integrated into the immediate diagnostic and treatment process.

Figure 1B shows the distribution of blood testing panels supported by POC devices. Clinical chemistry tests and blood glucose/diabetes monitoring dominated the panel with 26 devices (30.2%) each, reflecting the importance of these tests in chronic disease management, metabolic screening, and routine health monitoring. Complete blood count (CBC/FBC) came in second with 19 devices (22.1%), highlighting its ubiquity and necessity in daily clinical diagnosis. There were significantly fewer panels of more specialized tests such as HbA1c (6 devices, 7.0%), critical care and coagulation (3 devices, 3.5%), specific biomarkers (3 devices, 3.5%), hemoglobin (2 devices, 2.3%) and infectious diseases (1 device, 1.2%). This distribution may reflect the high technical barriers to the types of tests mentioned above, or a relative lack of market demand.

The 2D cross-frequency matrix heat map (Figure 1C) further revealed specific correlations between technology platforms and blood test types. A significant correlation between different technology platforms and test panel types was confirmed by χ^2^ test (χ^2^ = 237.81, df = 49, P < 0.001, Cramér’s V = 0.37). Electrochemical biosensor technologies were highly concentrated in blood glucose/diabetes testing, accounting for 64.0% of the technologies in this category, highlighting their dominance in the field of diabetes management. Microfluidic platforms were mainly used in clinical chemistry testing, accounting for 84.6% of the total, reflecting their prominence in multiple biochemical analyses. Flow cytometry technology was almost exclusively applied to CBC/FBC, demonstrating the technology’s specialized advantages in cell counting. In addition, fluorescence/immunoassay technology is more frequently applied to special biomarkers and infectious disease detection, reflecting its potential in the field of specialist diagnostics. Spectro-optical technology platforms are widely used in multiple types of tests such as clinical chemistry, blood glucose and HbA1c, demonstrating their adaptability in multiple testing scenarios. In contrast, imaging and microscopy technologies are only used for CBC/FBC, indicating their limited diagnostic applications.

In summary, there is a clear diagnostic application orientation to the choice of POC blood testing technologies. Electrochemical biosensors and microfluidic platforms excel in high-frequency, chronic disease management, while complex technologies such as fluorescence/immunoassay and flow cytometry are suited to more specialized diagnostic needs. At the same time, some key diagnostic areas such as infectious disease and critical care testing are significantly underrepresented in the current distribution of technologies, highlighting potential market and technology gaps for future research and development.

### 3.2 Time and Resources to Market by Medical Risk Class

This section synthesizes the impact of the regulatory pathway on the market entry of POC blood testing devices by analyzing the status of device regulatory approvals, the need for financing at different risk levels, and the average time required for regulatory approvals (Figures 1D and Figure 2A-B).

Figure 1D illustrates the distribution of the regulatory approval status of devices with respect to each regulatory category. Of the 86 devices analyzed, 88.4% have received at least one form of regulatory approval, while 11.6% have not yet received or have not disclosed their approval status. Of the approved devices, FDA Class II devices dominate (44.2%), followed by IVDR Class C devices (27.4%), indicating that medium-risk devices are most prevalent in the current market. Meanwhile, higher-risk regulatory categories such as IVDR Class D, MDR Class IIB and NMPA Class III account for a relatively small share of the market, at around 4-5 per cent each, reflecting the more stringent clinical evidence and higher cost inputs required for higher-risk regulatory pathways, potentially limiting the market entry of related devices.

Figure 2A further explores the capital requirements of equipment with different regulatory risk levels at different stages of financing from an economic input perspective. After dividing the equipment according to the risk level (low, medium, and high risk), it is found that the capital requirements are clearly and positively correlated with the risk level. Low-risk devices have low financing needs ($1.8 million on average) in the early stages (Early stage), with capital needs rising rapidly as the risk escalates to Medium and High risk, Medium-risk devices reach an average of $143.8 million at the Venture stage, and High-risk devices reach a maximum average of $126.5 million at the IPO stage.This trend clearly indicates that the capital requirement for devices increases significantly as the regulatory risk level increases, reflecting the fact that high-risk devices require greater financial investment during the clinical validation, regulatory approval and scale-up phases. The significance of this difference was further confirmed by Kruskal-Wallis test analysis (χ^2^ = 12.75, df = 2, P = 0.002), with capital requirements for medium- and high-risk devices being significantly higher than those for low-risk devices, reinforcing the strong link between regulatory risk and financing needs.

Figure 2B illustrates the distribution of approval times required for devices under different regulatory categories. Overall, the regulatory approval time increases significantly as the risk level of the device increases. The average approval time for FDA Class I devices is very short (about 7 days) due to the exemption of pre-market approval. the approval time for FDA Class II and III devices is 177 days and 243 days, respectively; and the approval time for China’s NMPA Class II and III devices is 180 days and 300 days, respectively, which is relatively efficient. FDA Class II and III devices take 177 days and 243 days, respectively; China’s NMPA Class II and Class III devices take 180 days and 300 days, respectively, which is relatively efficient. Under the European regulatory regimes (MDR and IVDR), the time taken for approval is longer and increases significantly with the level of risk, with the average approval time for MDR Class IIa and IIb being around 300-360 days, while the average approval time for IVDR-regulated Class B, C, and D devices reaches 365, 450, and 540 days, respectively. This on top of the R&D development which is calculated to be 10 years on average per device, independent of class. This variation across regulatory systems reflects differences in the stringency of regulatory approvals across regions, with longer approval times especially in the high-risk in vitro diagnostic space.

### 3.3 Usability in Near-Home Settings

This section evaluates the usability of POC blood testing devices in Near-Home Settings, including the distribution of device use environments (Fig. 2C), affordability (Fig. 2D), and physical usability (device volume and weight, See Supplementary Figures), with the aim of providing an empirical basis for device development for both home and non-specialist healthcare environments.

Figure 2C presents the distribution of the 86 analyzed devices in different usage scenarios. Of these devices, 51.9% are specifically designed for Point-of-Care (PoC) scenarios, 42.3% are for home use, and only 5.8% combine both home and PoC scenarios. This distribution clearly shows that although Point-of-Care scenarios still dominate, the proportion of home-use devices has gradually become significant, reflecting the trend of the POC market gradually shifting towards home health management, telemedicine and remote digital care.

Figure 2D shows the difference in economic costs between home and instant access devices. The results show that devices designed for home use are significantly lower in both purchase price ($590 on average) and annual cost of use ($458 on average) than just-in-time diagnostic devices ($1,637 purchase price and $1,435 annual cost of use). This difference clearly reflects the strong link between device cost and usage scenarios, with home devices being more cost-sensitive to accommodate the affordability of lay user needs. To assess physical usability, we further analyze the distribution of device volume (Figure S1) and weight (Figure S2) (See Suppl. Info). Analysis by the Mann-Whitney U test showed that there were significant differences between home-use devices and just-in-time diagnostic devices in both volume (U = 372, P < 0.001) and weight (U = 315, P < 0.001):

Thereby, the current wearable device market exhibits two clear trends: the high prevalence of long-term data storage capabilities and the rapid but still insufficient integration of AI capabilities. This suggests that future device development will need to focus on increasing the analytical value and actionability of user data while further strengthening the integration of intelligent algorithms. If wearable devices can effectively integrate continuous data collection and advanced intelligent analysis functions, they will certainly play a greater role in chronic disease management, personalized medicine and health monitoring, reflecting higher medical and market value.

### 3.5 Deployment Capabilities of POC Device Types

This section analyses the two key dimensions of economic burden (including the initial price of the equipment and the cost of consumables) and actual operational capability (training requirements, maintenance complexity, and system integration capability) embodied in different types of POC blood testing equipment in the actual deployment process, in order to gain a deeper understanding of the suitability of the equipment types for the actual deployment scenario (Figure 3A-B).

Figure 3A presents the average of the initial purchase price of the equipment versus the annual consumable usage cost for the four main POC equipment types. The results show that the Benchtop Hematology Analyzer (Benchtop Hematology Analyzer) has the highest initial purchase price ($6,885 on average), while the Benchtop Chemistry Analyzer (Benchtop Chemistry Analyzer) has the highest annual consumable cost ($6,514). In comparison, the Handheld POC Analyzer was in the middle of the pack in terms of both initial device cost ($2,644) and consumable cost ($2,592); the Microfluidic Cartridge/Chip Analyzer had the lowest initial device price ($698) but was not as expensive as the Benchtop Chemistry Analyzer due to consumable costs. The Microfluidic Cartridge/Chip Analyzer had the lowest initial equipment price ($698), but due to the high consumption of consumables, the annual cost of use was $2,346, which significantly exceeded the initial investment.

To further confirm the statistical significance of the above cost differences, the Kruskal-Wallis test was used in this study to analyze the difference in consumable costs, and the results showed that there was a significant difference in the cost of consumables between the different equipment types (χ^2^ = 9.34, df = 3, P = 0.025), with desktop equipment having a significantly higher cost of consumables than portable equipment.

Analysis for the practical operational capability dimension (Figure 3B) showed that the handheld instant diagnostic analyzer performed best in the deployment capability composite score (composite score of 4.48), with low training difficulty (4.58) and high system integration capability (4.85), reflecting high user-friendliness and system compatibility. The benchtop chemistry and hematology analyzers, while also having high system integration capabilities (4.89 and 4.47 respectively), have relatively low training and maintenance scores (around 3) due to their high technical complexity, limiting their widespread use in non-specialist and low-resource environments. Microfluidic chip analyzers had the lowest system integration score (3.40) and relatively lowest overall deployment score (3.33), suggesting that such devices face significant compatibility and integration challenges when deployed.

### 3.6 Global Distribution of POC Device Innovators

To further understand the global innovation landscape in the field of POC blood testing equipment, this study analyses the geographical distribution of companies engaged in the research and development of POC equipment on a global scale, as shown in Figure 3C. The results show that POC device innovators show a clear geographical concentration globally, with the United States, China, and some European countries leading the way.

The United States was found to be the most important global innovation center for POC blood testing devices as it is for better or worse, the most attractive market, with the largest number of device manufacturers, reflecting strong R&D capabilities and a highly active medical technology market despite its many healthcare system challenges, specially its lack of universality and affordability. This is closely followed by China, where the number of POC device manufacturers has increased rapidly in recent years, reflecting a rapid expansion in technological advances in the country’s medical device industry. Europe, on the other hand, has Germany, France and the UK as the main innovation clusters, demonstrating the region’s significant strengths in advanced medical technology research and development.

Notably, other regions such as Israel and Canada show some degree of innovation dynamism. This uneven global distribution suggests that future POC device R&D and healthcare resource allocation still needs to focus on geographic balance to enhance global healthcare equity and accessibility.

### 3.7 Continuous Monitoring, at Home Use capabilities and Remote Smart Testing

Home-use devices were smaller, with most devices having a volume of less than 1 dm³ (Fig. 4), whereas just-in-time diagnostic devices were significantly larger, with a median volume of approximately 10 dm³. Similarly, in terms of weight (Fig. 4), the home devices were significantly lighter, with the majority weighing less than 500 g, whereas the just-in-time devices clustered in the 2,000 to 10,000 g range and contained several heavier models that were outside the typical range.

**Figure 4:**
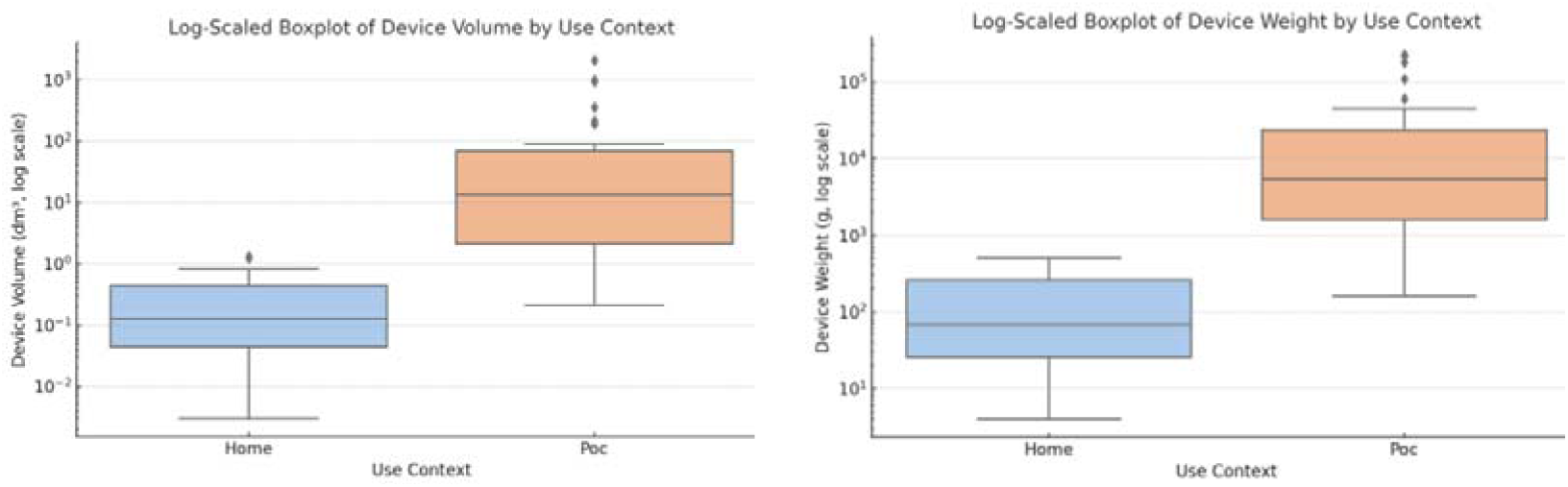
Log-Scaled Comparison of Device Volume and Weight by Use Context

These differences clearly reflect a greater emphasis on lightweight and compactness in the design of home-use equipment to enhance user-friendliness and portability in non-professional use environments. Immediate diagnostic equipment, on the other hand, although it may have advantages in terms of analytical performance and detection capability, its larger size and weight limit its use in non-professional, portability-demanding home environments.

In summary, the results of the analyses in this section show that the devices show a clear cost and physical attribute orientation in the near-home scenario. Home-based devices are designed with a focus on low cost, small size and low weight, and are more suitable for daily self-health management by non-professional users. Immediate diagnostic devices, on the other hand, focus more on diagnostic performance and analytical capabilities, and are usually dominated by the usage needs of professional medical scenarios. These findings clearly indicate that the development of devices for home scenarios needs to further focus on affordability, portability and user experience design to better meet the rapidly growing needs of the home healthcare market.

Figure 6 illustrates the distribution of different device types in terms of the combination of AI and long-term data storage features. Overall, the Handheld POC Analyzer and Benchtop Chemistry Analyzer lead in the number of devices with both AI and long-term data storage capabilities, demonstrating the combined strength of these two types of devices in intelligent data analysis and trend monitoring. Non-invasive glucose monitoring devices (Non-invasive Glucose Monitor) and benchtop hematology analyzers (Benchtop Hematology Analyzer) also showed high penetration of long-term data storage capabilities, but relatively low AI integration. Continuous CGM and Microfluidic Cartridge/Chip Analyzer show lower overall numbers, indicating that there is still more room for development of these devices with both AI and long-term storage. It is worth noting that the Sample Collection Device and Lateral-Flow/Rapid Test are almost devoid of these two advanced functions, highlighting the obvious functional gap between different device types in terms of data processing and intelligent analysis capabilities.

**Figure 5:**
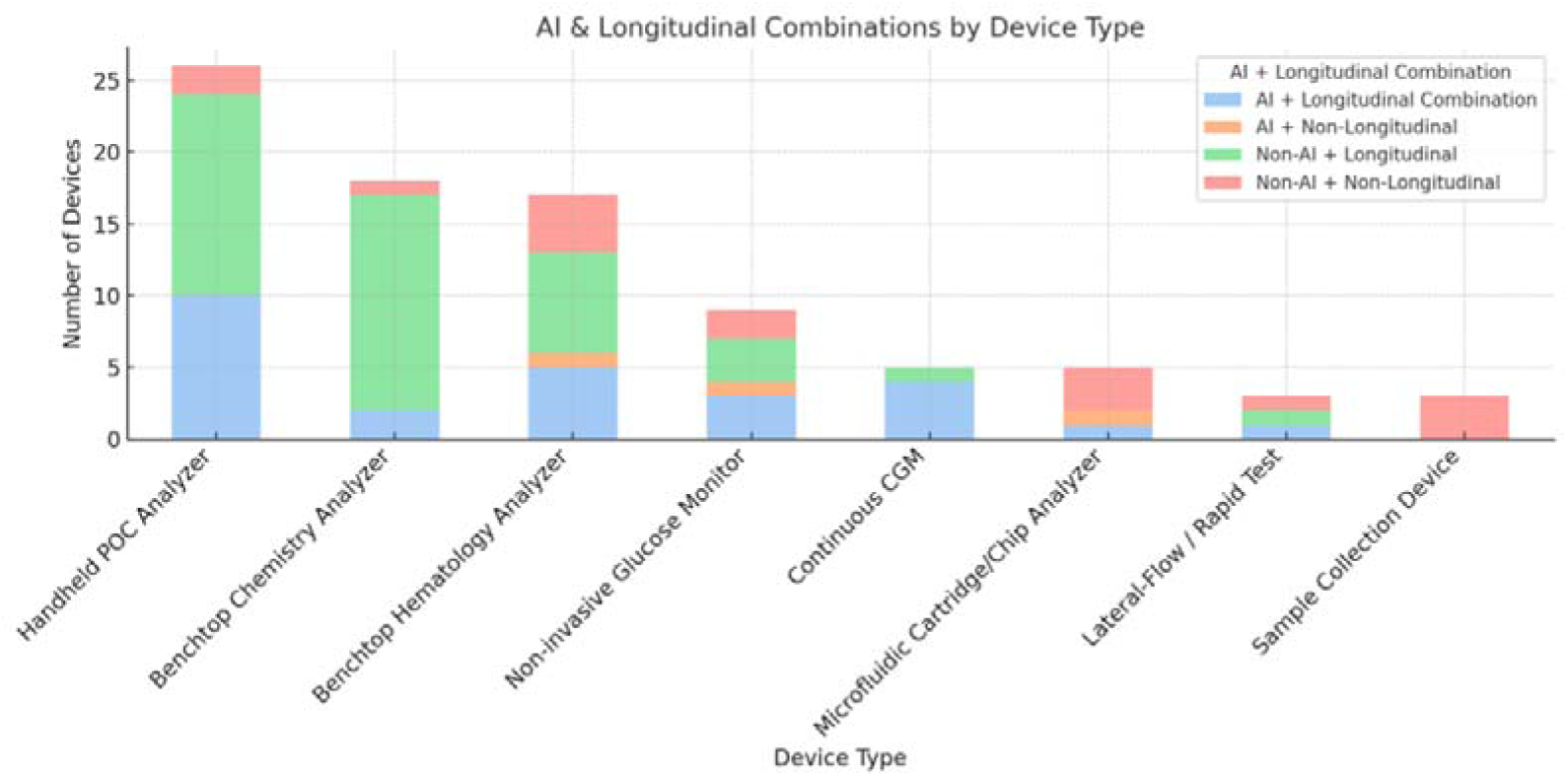
AI and Longitudinal Data Capabilities Across Device Types

**Figure 6:**
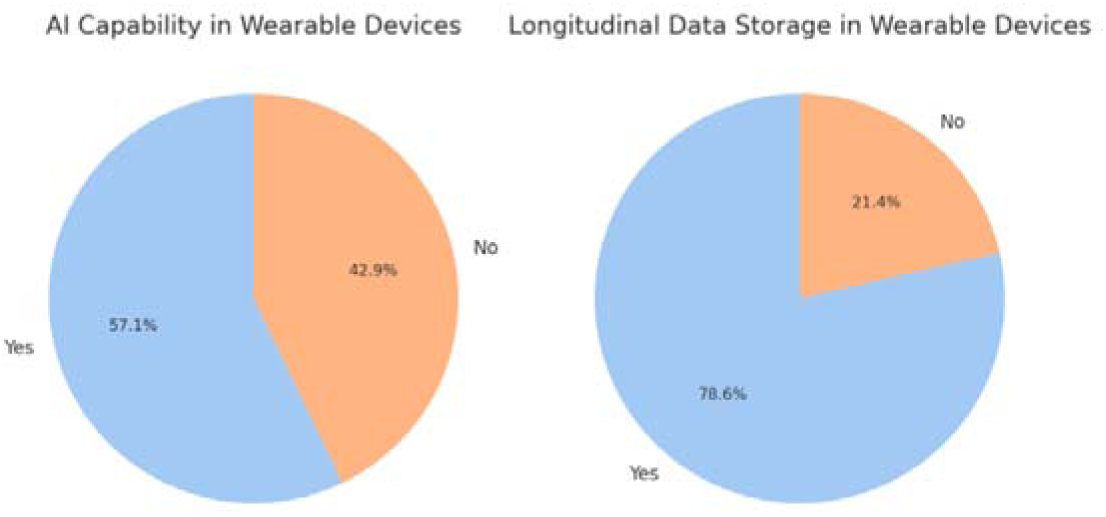
AI and Longitudinal Data Capabilities in Wearable Devices: While POC devices tend to lack AI and continuous data collection capabilities. At-home and wearables

Figure 6 clearly shows that 57.1% of wearable devices have AI capabilities, implying that these devices are progressively integrating intelligent analytics for more personalized and real-time health management services. However, there are still 42.9% of wearables that are not yet AI-capable, indicating that the progress of intelligent integration in the market still needs to be further improved.

Figure 6 shows that the penetration rate of long-term data storage in wearable devices reached 78.6%, significantly higher than the penetration rate of AI functions, indicating that device developers are more inclined to satisfy users’ needs for long-term trend analysis and data monitoring first, so as to lay the foundation for continuous health monitoring before gradually introducing more complex AI analysis functions.

## 4. DISCUSSION

Five themes frame the contemporary POC diagnostics landscape: (i) electrochemical biosensing as the leading modality, (ii) risk-adjusted regulatory pathways shaping time-to-market and cost, (iii) distinct usability and safety envelopes for home vs. professional use, (iv) the growing but selective role of longitudinal data and AI in wearables, and (v) pragmatic deployment trade-offs between capability and simplicity. These converge with prior literature while adding quantitative nuance from our device-level mapping.

Electrochemical biosensors underpin nearly one-third of devices (Sec. 3.1), consistent with their sensitivity, low cost, and integration into strips/cartridges, i.e., factors repeatedly cited as decisive for translation [99–101]. Our panel distribution, dominated by glucose and chemistry assays, mirrors market syntheses listing glucose, pregnancy, fecal occult blood, strep antigen, and coagulation as most common POC offerings [101]. Advanced modalities such as optics and microfluidics remain less represented due to cost and maturity constraints.

Regulatory analysis (Sec. 3.2) shows higher-risk or novel devices incur longer review times and financing needs, paralleling evidence from U.S. and EU pathways [102,103]. This gradient aligns with FDA’s 510(k)/De Novo/PMA distinctions and EU MDR Rule 11, confirming how regulatory burden channels development strategies.

Near-home devices differ structurally from professional POC tools, as reflected in FDA usability guidance [104] and IEC 60601-1-11 [105]: emphasis lies on lay-safety, robustness, and low training burden. In wearables (Sec. 3.4), connectivity and longitudinal logging diffuse faster than validated ML decision support, echoing reviews highlighting the gap between continuous data and outcome-linked AI [106]. Deployment patterns (Sec. 3.5) reflect ASSURED/REASSURED criteria [3]: handhelds dominate where simplicity matters, benchtops trade cost for breadth, and microfluidics balance capability with consumable and IT burdens [107,108].

Collectively, our findings reinforce electrochemistry as today’s workhorse, regulatory risk as a driver of cost/time, user-safety as central for home POC, data/AI integration as emergent, and deployment choices as shaped by pragmatic trade-offs, i.e., features consistently mirrored in the wider POC ecosystem.

### 4.2 Limitations

No review can claim to be complete. This study included 86 devices, capturing major global portfolios but not all innovations, especially in LMICs. Reliance on public specifications and filings means some features may reflect claims rather than validated performance. Scoring involved some subjectivity despite structured criteria. Absence of clinical outcome data limits generalizability: device favourability in analysis may not equate to patient benefit, a gap emphasized by Mitra & Sharma (2021) [110]. Finally, the cross-sectional design represents a 2025 snapshot in a rapidly evolving field.

### 4.3 Future Research Directions

Future work should:

1. **Expand regional coverage:** Include LMIC-developed devices, which often prioritize frugal, durable innovation [110,111].
2. **Evaluate real-world usability and outcomes:** Conduct formal usability studies and clinical trials to test impacts on care decisions and patient outcomes [112].
3. **Address sustainability:** Perform lifecycle assessments to reduce e-waste and carbon impact; explore biodegradable and recyclable components [113].
4. **Advance AI integration:** Develop and validate AI-enabled continuous monitoring under transparent governance, building evidence on safety and user trust [106,114].
5. **Explore systemic issues:** Standardize data integration, enhance training/education, and examine reimbursement/policy frameworks to support equitable adoption.
6. **Different weighting options:** In a setting of smart remote care, size, data collection or AI capabilities could play a higher role and weight more in the raking index.
7. **Early-stage R&D:** The study could include devices currently under development.
8. **Integration to current pathways and adoption:** How easy is to integrate to current EMR, EPR and LIMS systems, and how easy or affordable its adoption in actual healthcare patways may be.
9. **Causal Intervention:** Finally, future work should explore causal discovery and intervention modelling using *algorithmic information dynamics (AID)* to identify minimal, structure-altering perturbations and intervention points in diagnostic ecosystems. AID provides a principled framework for mapping causal mechanisms, reconstructing generative rules, and testing counterfactual intervention strategies in complex biomedical systems [115–118]. Integrating these methods with POC longitudinal data streams may reveal hidden causal drivers of diagnostic performance, device behaviour, or patient-level physiology.

Widening geographic scope, intended use, validation of clinical outcomes, sustainability, affordability, and embedding AI will be critical to ensure POC, portable and wearable devices are clinically impactful, environmentally responsible, and equitably accessible worldwide.

## 5. CONCLUSIONS

This study provides a comprehensive analysis of innovation in point-of-care (POC), portable and wearable blood testing technology, evaluating 86 products across technological platform, diagnostic scope, regulatory pathway, cost, usability, and global distribution. Key trends include the predominance of portable electrochemical biosensors for routine assays (notably glucose), while more complex modalities remain niche due to cost and specialized requirements. The diagnostic menu is broad but uneven: metabolic panels and blood counts are well represented, whereas critical care and infectious disease markers have relatively few POC options. Most devices fall into moderate-risk regulatory classes that allow shorter, less burdensome approvals; high-risk novel devices are rare, constrained by longer timelines and evidence requirements. Usability and cost show clear trade-offs: home-use devices emphasize affordability and simplicity, while clinical systems deliver broader capabilities at greater cost and complexity. Connectivity is now common, but fully AI-integrated diagnostics remain limited to a few platforms. Innovation is geographically concentrated in North America, Europe, and East Asia, revealing disparities in access.

The 86-device sample may underrepresent new or region-specific innovations, and reliance on proxy measures (advertised features, regulatory status) introduces potential bias. Devices were not validated in clinical practice, so favorable lab profiles may not translate to patient benefit. Rapid innovation means our results reflect a snapshot rather than a forecast. Broader risks also emerge ensuring that POC devices improve outcomes will require rigorous validation, user training, and sustainable design to mitigate e-waste. Commercial adoption will depend on cost-effectiveness and supportive reimbursement, especially in low-resource settings.

Future research should expand coverage to LMICs and underrepresented regions, evaluate devices in real clinical workflows, and incorporate sustainability through greener manufacturing, reusable materials, and waste management. Advancing AI and continuous monitoring must be paired with rigorous validation to ensure trust, privacy, and preventive-care value. Finally, cross-disciplinary collaboration among engineers, clinicians, regulators, and economists will be essential to tackle training, health IT integration, and business models. By addressing these priorities, POC, portable and wearable diagnostics can move from promising prototypes to safe, effective, and widely accessible tools for global health.

## Data Availability

All data analyzed in this study were derived from publicly available sources, including regulatory databases (e.g., FDA, IVDR, NMPA), manufacturer technical documentation and Instructions for Use (IFUs), academic literature, and commercial intelligence platforms cited in the manuscript. The curated dataset, classification framework, and derived summary tables generated during the analysis are contained within the manuscript and its supplementary materials. Additional processed datasets supporting the findings of this study are available from the corresponding author upon reasonable request.

## Notes

### Competing Interest Statement

The authors have declared no competing interest.

### Funding Statement

There is no public data availability link; the data are compiled from publicly available sources and are available from the corresponding author upon reasonable request.

